# Making sleep behaviors interpretable: adapting the two-process model of sleep regulation to longitudinal Fitbit sleep and activity behaviors for health insights

**DOI:** 10.64898/2026.03.01.26347356

**Authors:** Peyton L. Coleman, Jeffrey Annis, Hiral Master, Daniel E. Gustavson, Lide Han, Evan Brittain, Douglas M. Ruderfer

## Abstract

**Background:** As sleep data from wearable devices are increasingly available in health research, there are new opportunities to understand sleep regulation behaviors as modifiable risk factors for disease. At such a large scale (tens of thousands of people over millions of day-level observations), prioritizing and interpreting sleep behaviors is challenging while maintaining biological relevance and modifiability. In this work, we aim to address this challenge by proposing a framework to interpret Fitbit data through a well-known neurobiological framing of sleep regulation, the two-process model.

**Methods:** We use data from the All of Us Research Program, a national biobank with passively collected Fitbit data for 32,292 people across 15,754,893 total days. We map Fitbit behaviors (_b_) to either circadian (C) or homeostatic (S) processes. Using iterative exploratory factor analysis to obtain weights, the Fitbit C_b_ and S_b_ are then weighted at the level of each day to create C_b_ and S_b_ scores.

**Findings:** C_b_ and S_b_ scores were found to align with expected real-world relationships with age, seasonality, shift work, and napping. C_b_ and S_b_ scores were interpreted with relation to depression, where it was found that S_b_ scores are highly associated with likelihood of diagnosis (OR = 1.5, p < 2e-16) while C_b_ and S_b_ scores are equally associated with severity (S_b_ score β = 0.2, C_b_ score β = 0.21, p < 2e-16).

**Interpretation:** C_b_ and S_b_ scores support longitudinal interpretation (e.g., changes in S_b_ around treatment), aggregation (e.g., differences in C_b_ between two groups), and actionable modification (e.g., reduce naps to improve poor S_b_). Overall, our behavior scores allow for interpretation of wearables sleep data and can be utilized across many disease contexts to better understand how sleep influences health.

**Funding:** This work was supported by NIH training grant T32GM145734 and NIH R21HL172038.

## Introduction

Rapid technological advancement in wearable devices (e.g., Fitbits, Apple Watch) has enabled high resolution capture of behavior at scale. Wearables data has been collected in tens of thousands of participants and integrated with extensive clinical data in population biobanks to understand the impact of behavior on health.^1^ These data are particularly relevant for studying sleep, as capturing longitudinal data on sleep at scale has been historically difficult.^2^ Electroencephalography (EEG) and polysomnography are the gold standard for sleep measurement but require in-lab sleep sessions and obstructive measurement devices that are not practical for extensive longitudinal assessments. Repeated self-reported sleep surveys are scalable but lack reliability.^3^ Wearable derived sleep measures have shown validity and scalability presenting an opportunity to improve our understanding of how sleep affects health and vice versa.^4^

An important component to this goal is maintaining the interpretability of sleep behaviors, particularly relating these behaviors to processes that could implicate possible treatments and/or those that might be modifiable. One way to add interpretability is by relating sleep behaviors back to their biological frameworks. Fortunately, sleep regulation has been thoroughly modeled in neuroscience research which we can harness to interpret sleep behaviors. The two-process model of sleep regulation, first posited in 1982, explains sleep regulation as a combination of a circadian (C) and a homeostatic (S) process, which together influence timing, duration, and quality of sleep.^5^ Process C (**Figure 1A**) can be thought of as the body’s internal clock. It operates closely to a daily 24-hour cycle and is largely influenced by external stimuli, such as sunlight in the morning and darkness in the evening, which then results in the body waking and sleeping in time with the earth’s daily rotation.^6^ Process C is largely modulated by melatonin in the suprachiasmatic nucleus. By contrast, process S captures sleep pressure built up since the last sleep period. In other words, the longer you are awake, and the more active you are, the more tired you become (**Figure 1A**).

**Figure 1.**
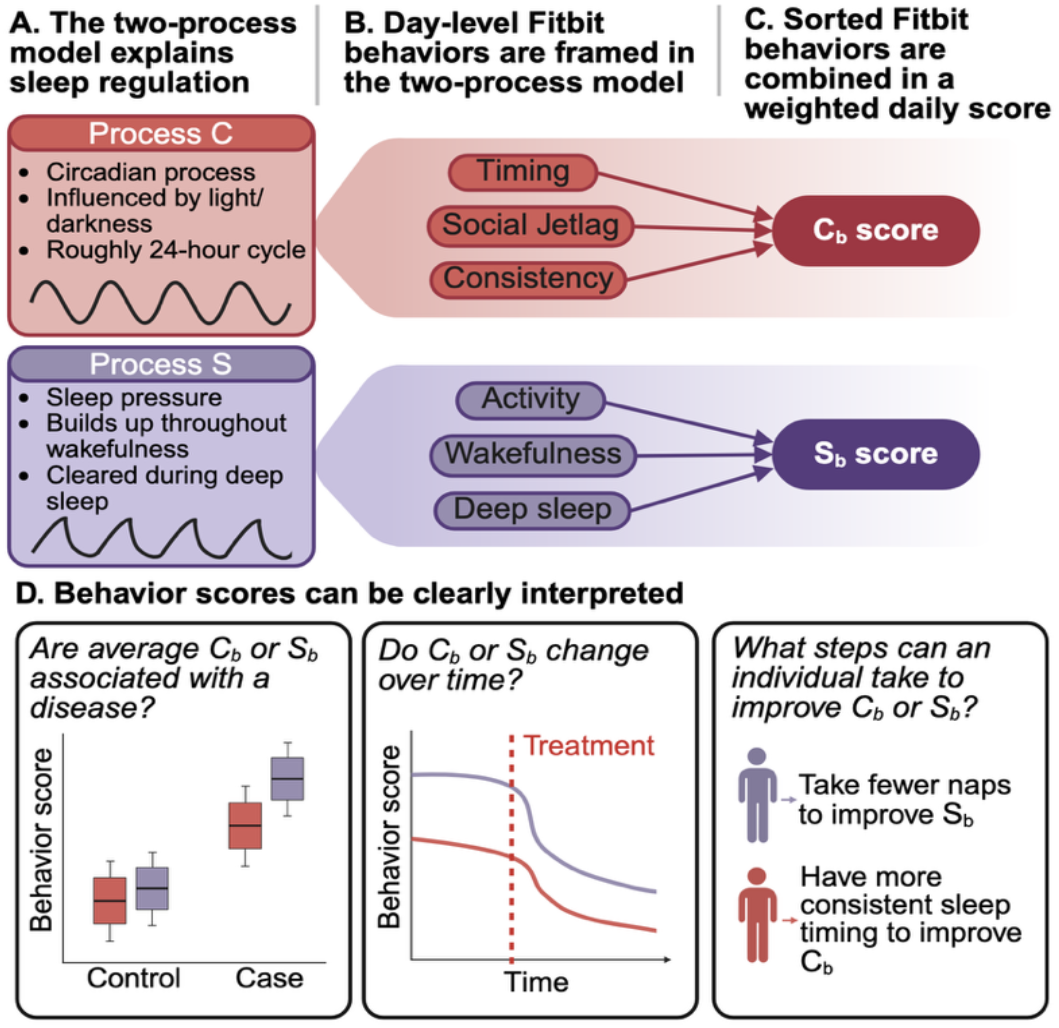
Framework to interpret Fitbit behaviors through the two-process model of sleep regulation. **(A)** The two-process model of sleep regulation explains sleep regulation as the combination of circadian (C) and homeostatic (S) processes, which are independent and sufficient to induce sleep but interact with each other. **(B)** We have curated and mapped Fitbit-measured behaviors (_b_) to either process C or S based on literature and previous studies of sleep systems. **(C)** The mapped Fitbit behaviors are weighted using factor scoring to create a C_b_ and S_b_ score for each day. **(D)** Example interpretations of C_b_ and S_b_ scores.

Neurobiologically, process S is explained by the buildup of adenosine in the brain as a byproduct of cellular metabolism and activity, which causes tiredness by inducing hyperpolarization in “wakefulness” cholinergic, serotonergic, and noradrenergic neurons to disinhibit the hypnogenic system. Once sleep begins, the sleep pressure is relieved, and the buildup begins again once you wake up. Crucially, the two processes have different implications for behavioral intervention: process C disruptions may be treated with melatonin supplement, routine changes in sleep/wake timing, or light therapies, while process S disruptions may be more likely to respond to increased activity, sleep restriction therapy, or SSRI/SNRIs.

The two-process model is ideal for framing sleep behaviors because it allows for clear interpretation. As an example, depression has been associated with both less and more average sleep duration, higher variability in sleep duration and shorter average deep sleep segments.^7^ Depression is also comorbid with both insomnia and hypersomnia, which at first glance appears to be contradictory and difficult to interpret. However, these sleep issues are primarily due to a deficiency of process S, due to lessened activity and serotonergic disruption as core traits of the disorder.^8,9^ This process S deficiency then causes issues sleeping at night. In this case, antidepressants or increased daytime activity may be effective interventions.

This paper will present a framing of longitudinal Fitbit sleep behaviors based on the two-process model of sleep regulation. We map Fitbit-derived behaviors (_b_) to circadian (C) and homeostatic (S) processes (**Figure 1B**), construct weighted C_b_ and S_b_ scores (**Figure 1C**), and evaluate their credibility and clinical interpretability in the All of Us Research Program.^1^ The benefit of framing sleep-related behaviors with the two-process model in health research is twofold. First, this framing allows for more clear and clinically actionable recommendations to promote better sleep regulation (**Figure 1D**). For example, if we know a particular person has high process S behavior disruption, we may be able to recommend process S-encouraging behaviors, such as refraining from naps. Second, since the two-process model describes underlying biological processes, by framing behaviors in this way we may be able to glean insights into the biology of sleep regulation for the first time at scale.

## METHODS

### Cohort Description

We analyzed data from the All of Us Research Program (AoURP) version 8 where electronic health record data and passively collected Fitbit data existed for 32,392 individuals.^1^ Fitbit data came from two sources: (1) Bring your own device program (BYOD) - participant-volunteered devices with retrospective monitoring periods (n = 13,048), and (2) the WEAR sub-study, which provides Fitbit devices to a diverse cohort at no cost with approximately one year of monitoring (n = 14,263). We analyzed both cohorts together and adjusted for cohort in sensitivity analyses (**Supplemental Fig 1**).

### Fitbit Data Quality Control

First, we processed sleep data through a previously published “typical sleep period” (TSP) cleaning algorithm.^10^ Briefly, the TSP uses an individual’s median sleep times to consolidate sleep fragments into a main sleep periods (e.g., combines a sleep segment from 11:00PM-1:30AM and 3:30AM-7:00 AM). This pipeline more accurately identifies sleep periods with large awakening periods. We restricted to days with less than 1,000 minutes (16.6 hours) of sleep duration and a minimum of 14 days per participant, resulting in 15,741,655 valid total days across 32,386 individuals (**Supplemental Fig 2**).

### Feature Engineering

Our goal was to map behaviors to process C (C_b_) and S (S_b_) and calculate weighted behavioral scores for each day, which we will refer to as C_b_ and S_b_ scores. We compiled several Fitbit-measurable behaviors related to C and S based on prior literature and expert review (**Supplemental Fig 3**). These behaviors (C_b_ and S_b_) were then combined into day-level scores, detailed later.

### C_b_ (circadian behaviors)

#### Phasing-related behaviors

To capture circadian phasing, we utilized bed and wake timing (clock time) for each typical sleep episode. Additionally, ZIP code information was used to ascertain sunset and sunrise for each date using the bioRad R package, which we then used to calculate absolute differences between sunset-bedtime and sunrise-wake time.

#### Consistency-related behaviors

To capture circadian consistency, we calculated the absolute difference in midsleep point (halfway between bedtime and waketime) between each night and the night prior. For social jetlag, a common metric used to identify circadian misalignment,^11^ we calculated the absolute difference between a day’s midsleep and the median midsleep of the prior two weekend days (if weekday) or five weekdays (if weekend).

#### C_b_-specific Quality Control

Engineering of C_b_ required participants to have ZIP code information. We also required participants to have at least seven pairs of consecutive days to ascertain consistency. After this filtering, there are 9,947,327 days with valid C_b_ across 27,039 individuals.

### S_b_ (homeostatic behaviors)

#### Wakefulness-related behaviors

Since process S builds up during prolonged wakefulness, we calculated a metric (prior wakefulness) to capture the amount of wakefulness before the typical sleep episode. For most days, this would be the amount of time from the previous wake time (e.g., 16 hours if waketime is 7:00 AM and bedtime is 11:00 PM). On days with naps (defined as sleep outside the typical sleep period), prior wakefulness is reduced. For example, if waketime was 7:00 AM and bedtime was 11:00 PM, but there is a nap from 1:00 PM to 4:00 PM, the prior wakefulness for that day would be seven hours (4:00 PM – 11:00 PM). This metric accounts for the depletion of S buildup after naps.

#### Activity-related behaviors

Process S buildup is also influenced by activity during wakefulness, as activity increases adenosine in the brain and promotes tiredness. To capture daily activity, we utilized total steps per day and the peak ten-minute step count (the mean steps per minute during the ten minutes of highest step count) as a proxy for intensity.

#### Staging-related behaviors

The most prominent known correlate of adequate process S buildup is the pattern of sleep staging throughout the night.^6^ Deep sleep segments should be longer in the first half of sleep, and REM sleep segments should increase in length as sleep goes on. Despite known limitations of Fitbit sleep staging, notably an underestimation of deep sleep and an overestimation of REM sleep, we investigated whether there were Fitbit sleep staging features that accurately fit into this expected model. Through sensitivity analyses detailed in supplement, we identified two staging features that exhibited the correct directional association with naps and wakefulness: the length of the first deep sleep segment and the length of all deep sleep segments prior to the first REM segment. We also found that scaling (z-scoring) within the individual increased the association between these features and naps and wakefulness due to high individual differences in sleep staging (**Supplemental Fig 4**). These two features were thereby selected for use in subsequent analyses as process S-related sleep staging features.

#### S_b_-specific Quality Control

Outlier days with 1,000 minutes of sleep duration, 50,000 steps, six hours of fair or intense activity, 18 hours of light activity, and steps intensity of an average 300 steps-per-minute in a 10-minute period were removed. We required at least 85% of wakefulness to have confirmed wear time to ensure that prior wakefulness did not appear longer due to missingness during the daytime (**Supplemental Fig 5**). After this filtering, there are 8,210,714 days with valid S_b_ across 27,313 individuals.

### Factor Analysis and Scoring

We then combined the behaviors into weighted behavior scores. Our goal was to retain the relative weight of each behavior at the level of a single day, as opposed to taking means of behaviors over time. To accomplish this, we first standardized all the behaviors (z-scores) and restricted sampling days to only include behaviors within five standard deviations of the mean, to ensure weights were not overly influenced by outlier days. Next, we randomly sampled a single day per participant and used the sampled dataset in an exploratory factor analysis using promax rotation (**Figure 2A-B**). We then repeated this random sampling over 5,000 iterations to avoid within-subject correlation of behaviors (**Figure 2C**). Finally, we calculated the median loading for each behavior across iterations as its weight (**Figure 2D**). We oriented factor signs so a higher score represents worse C_b_ or S_b_. We then weighted each factor equally, summed the factor scores, and divided by the number of factors to get final C_b_ and S_b_ scores (**Figure 2E**).

**Figure 2:**
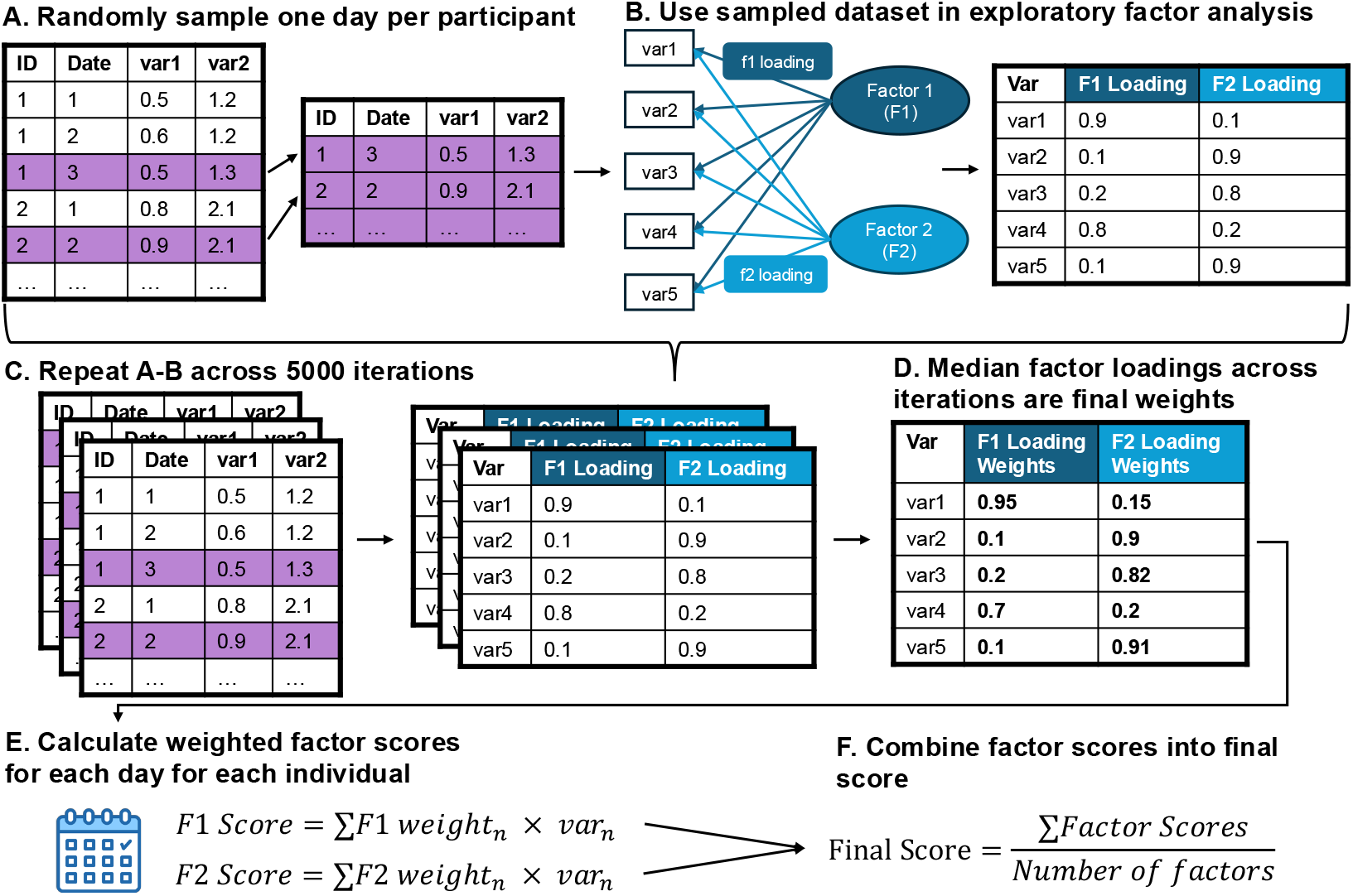
Schematic of factor scoring method to create day-level C_b_ and S_b_ scores. Days with missingness are excluded separately for C_b_ and S_b_. **(A)** A single day of behaviors is sampled from each participant. **(B)** Sampled days are modeled using exploratory factor analysis, and factor loadings for each behavior on each latent factor are collected. **(C)** A-B are repeated across 5,000 iterations of randomly sampled days. **(D)** From the 5,000 iterations, we calculated the median factor loadings of each behavior on each latent factor to serve as the factor score weights. **(E)** A weighted score is created for each latent factor by weighting each behavior with the median factor loading and then summing the weighted behaviors. **(F)** The final score for each day is created by summing the individual factor scores and dividing by the number of factors.

### Regression models of depression diagnosis and severity

To assess the clinical applicability of the C_b_ and S_b_ scores, we examined their relationships with depression, as the processes are both shown to be disrupted in depression, though C is thought to be less associated than S.^9^ Our definition of Major Depressive Disorder (MDD) diagnosis follows validated definitions that incorporate both EHR and survey data.^12^

#### EHR data

ICD (International Classification of Diseases) billing codes were mapped to PhecodeX, which groups billing codes into clinically meaningful diseases and conditions.^13,14^ The phecode for MDD (MB_286.2) includes ICD9 codes 292.2-292.26 and 292.3-292.36, as well as ICD10 codes F33.0-F33.4 and F33.8-F33.9.

#### Survey data

All of Us participants are eligible to complete an Emotional Health and Wellbeing survey, which includes two sub-surveys which can be used as an MDD diagnosis proxy. The CIDI-SF (Composite International Diagnostic Interview – Short Form)^15^ requires at least two weeks of anhedonia or dysphoric mood, as well as at least 5 depressive symptoms to be considered a diagnosis of depression. The PHQ-9 (Patient Health Questionnaire-9)^16^ asks a number of questions and returns a depression severity score. If the severity score was greater than 10, they are considered to have a diagnosis of depression.

#### Depression diagnosis

A diagnosis of MDD was defined by meeting at least two of the three following criteria: having at least two instances of an MDD phecode on different dates; one prescription for an antidepressant; a score of >10 on the PHQ-9 or qualifying for case status on the CIDI-SF, when available (**Supplemental Fig 6**).^12^ Antidepressants were identified using the National Drug File – Reference Terminology (NDF-RT) (**Supplemental Table 1**). Patients with a phecode for bipolar disorder (MB_286.1*), schizophrenia (MB_287.1), or schizoaffective disorder (MB_287.2) were excluded.

#### Regression models of depression

To model each item’s relationship with depression, each item was used as the independent variable in a logistic regression using MDD case/control status as the outcome, covarying for age, gender, race, and length of Fitbit recording. Additionally, to assess each item’s relationship with depression severity, we modeled PHQ-9 severity score using linear regression with the same covariates. As PHQ-9 scores are heavily skewed towards zero, we applied an ordered quantile transformation to the raw scores.

## RESULTS

### Cohort description

A total of 32,392 All of Us participants provided Fitbit sleep data linked to electronic health records (EHR) across 15,754,893 days before quality control (see Methods). After quality control, 27,313 participants remained, with most self-identifying as women (67.1%) and White (72.4%) (**Table 1**). The median age was 58.7 (range 21-100), and median monitoring length was 197 days (range 1-4,187 days). 18% of our cohort met criteria for a diagnosis of Major Depressive Disorder (n = 5,675), and 10,758 participants had completed at least one PHQ-9 survey.

**Table 1:** Cohort demographics.

|  | Count | Percent |
| --- | --- | --- |
| **Gender** |  |  |
| Woman | 18,341 | 67.2% |
| Man | 8,282 | 30.3% |
| Other | 690 | 2.5% |
| **Race** |  |  |
| White | 19,790 | 72.5% |
| Other | 2,198 | 8% |
| Black | 2,142 | 7.8% |
| More than one | 1,762 | 6.5% |
| Asian | 1,104 | 4% |
| Middle Eastern/North African | 160 | 0.6% |
| American Indian/Alaska Native | 157 | 0.6% |
| **Major Depressive Disorder** |  |  |
| Control | 19,045 | 77% |
| MDD Case | 5,675 | 23% |
| **Patient Health Questionnaire 9 Data** |  |  |
| No PHQ-9 | 17,150 | 62.8% |
| Has PHQ-9 | 10,163 | 37.2% |
| Median | SD | Range |
| **Age (years)** |  |  |
| 58.7 | 16.0 | 21 - 100 |
| **Fitbit observations (days)** |  |  |
| 218.0 | 700.0 | 1 - 4187 |

Quality control was applied separately for behaviors linked to process C (C_b_) and behaviors linked to process S (S_b_, see Methods). Briefly, for C_b_, we removed individuals without ZIP code information and without at least seven pairs of consecutive days. For S_b_, we removed days with extreme outliers of activity or sleep duration and required days to have at least 85% of wakefulness hours with confirmed waketime. After this quality control, there were 9,947,327 valid days with C_b_ across 27,039 individuals, and 8,210,714 valid days with S_b_ across 27,313 individuals.

**Table 2** displays the C_b_ and S_b_ descriptives across all days. The median wake time was 7:02 AM, and the median bedtime was 11:10 PM. The median number of steps was 6,635, and median wakefulness was 950 minutes (15.8 hours).

**Table 2:**
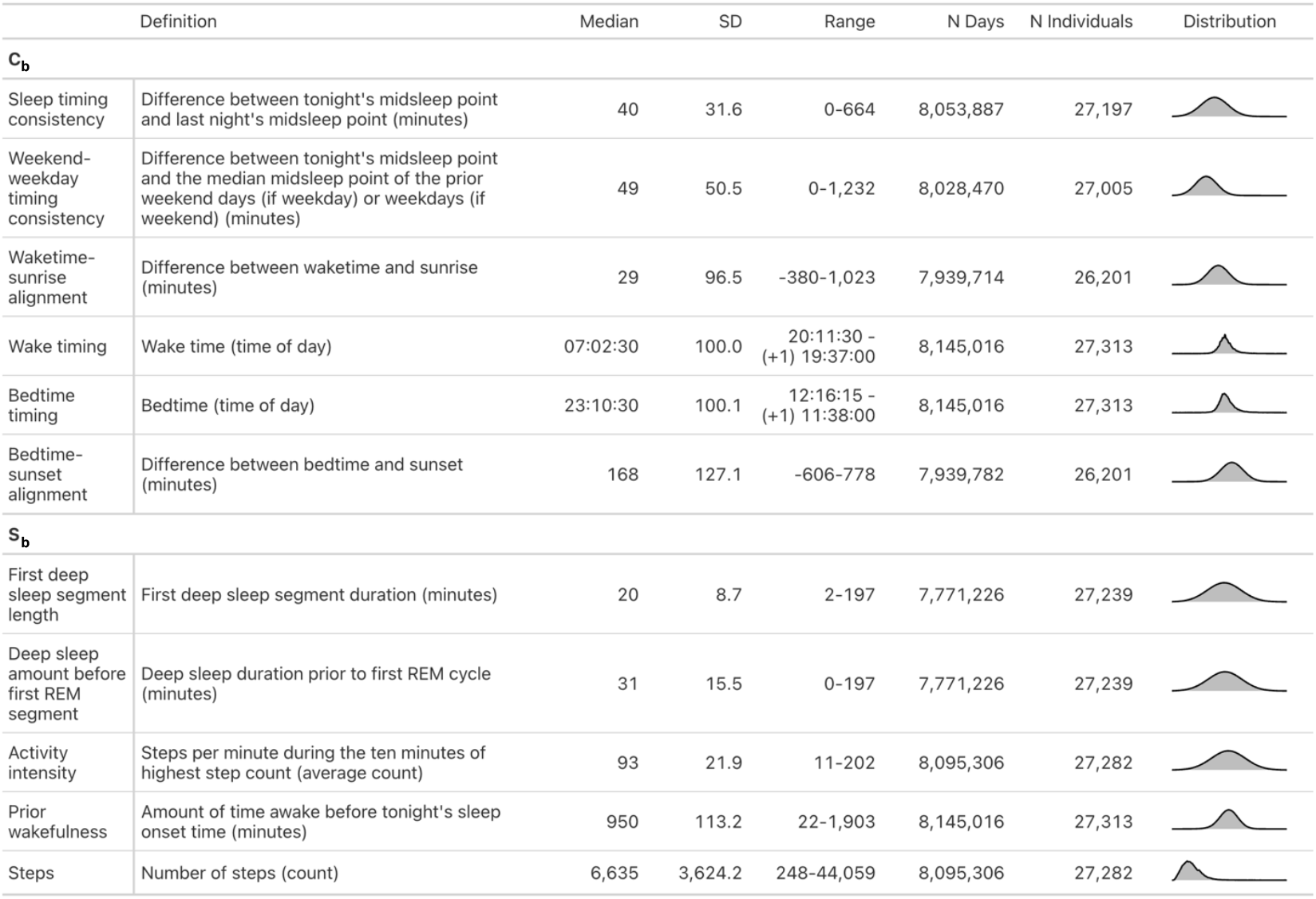
C_b_ and S_b_ definitions and descriptive statistics.

|  | Definition | Median | SD | Range | N Days | N Individuals | Distribution |
| --- | --- | --- | --- | --- | --- | --- | --- |
| **$C_b$** |  |  |  |  |  |  |  |
| Sleep timing consistency | Difference between tonight's midsleep point and last night's midsleep point (minutes) | 40 | 31.6 | 0-664 | 8,053,887 | 27,197 |  |
| Weekend-weekday timing consistency | Difference between tonight's midsleep point and the median midsleep point of the prior weekend days (if weekday) or weekdays (if weekend) (minutes) | 49 | 50.5 | 0-1,232 | 8,028,470 | 27,005 |  |
| Waketime-sunrise alignment | Difference between waketime and sunrise (minutes) | 29 | 96.5 | -380-1,023 | 7,939,714 | 26,201 |  |
| Wake timing | Wake time (time of day) | 07:02:30 | 100.0 | 20:11:30 - (+1) 19:37:00 | 8,145,016 | 27,313 |  |
| Bedtime timing | Bedtime (time of day) | 23:10:30 | 100.1 | 12:16:15 - (+1) 11:38:00 | 8,145,016 | 27,313 |  |
| Bedtime-sunset alignment | Difference between bedtime and sunset (minutes) | 168 | 127.1 | -606-778 | 7,939,782 | 26,201 |  |
| **$S_b$** |  |  |  |  |  |  |  |
| First deep sleep segment length | First deep sleep segment duration (minutes) | 20 | 8.7 | 2-197 | 7,771,226 | 27,239 |  |
| Deep sleep amount before first REM segment | Deep sleep duration prior to first REM cycle (minutes) | 31 | 15.5 | 0-197 | 7,771,226 | 27,239 |  |
| Activity intensity | Steps per minute during the ten minutes of highest step count (average count) | 93 | 21.9 | 11-202 | 8,095,306 | 27,282 |  |
| Prior wakefulness | Amount of time awake before tonight's sleep onset time (minutes) | 950 | 113.2 | 22-1,903 | 8,145,016 | 27,313 |  |
| Steps | Number of steps (count) | 6,635 | 3,624.2 | 248-44,059 | 8,095,306 | 27,282 |  |

### Modeling process scores for C_b_ and S_b_

To understand the relationships across the behaviors and define quantitative representations of the behaviors at the day-level, we utilized exploratory factor analysis for C_b_ and S_b_ across 5,000 random samples where each individual contributed data from a single day (**Figure 2**). Factor loadings for each behavior were then used as weights to create a C_b_ and S_b_ score model. Loadings greater than one were fixed to one.

#### Process C

C_b_ were best explained by a 3-factor model (**Fig 3A-C, Supplemental Fig 7**). Factor 1 had high loadings for consistency-related features (social jetlag 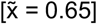 and night-to-night midsleep difference 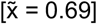); factor 2 had high loadings for bedtime-related features (bedtime 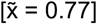 and bedtime-sunset alignment 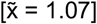); factor 3 had high loadings for waketime-related features (waketime 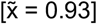 and waketime-sunrise alignment 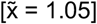. The final C_b_ scores weighted each behavior similarly, with bedtime timing weighted the highest [0.19], followed by bedtime-sunset alignment [0.17], midsleep point consistency [0.17], wake timing [0.16], waketime-sunrise alignment [0.16], and finally weekend-weekday timing consistency [0.15] (**Fig 2D**).

**Figure 3.**
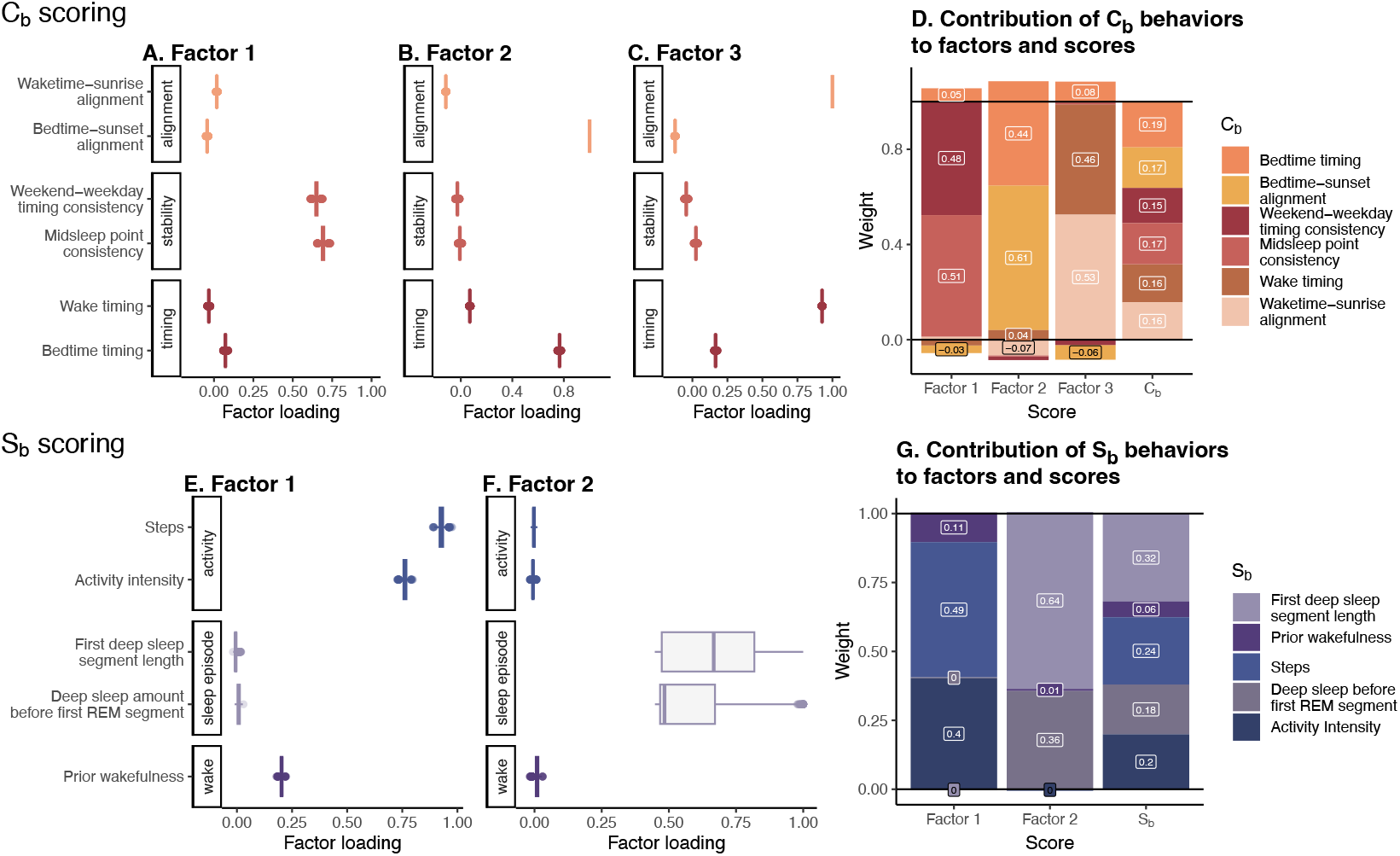
Determination of weights for C_b_ and S_b_ scores. **(A-C)** Factor loadings from exploratory factor analysis of day-level C_b_ onto three factors across 5,000 iterations. **(D)** Final median weight of each C_b_ for each individual factor and the final combined factor score. **(E-F)** Factor loadings from exploratory factor analysis of day-level S_b_ onto two factors across 5,000 iterations. **(G)** Final median weight of each S_b_ for each individual factor and the final combined factor score.

#### Process S

S_b_ were best explained by 2 factors (**Fig 2E-F, Supplemental Fig 7**). Factor 1 had high loadings for activity-related features (steps 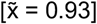 and steps intensity 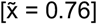) and moderate loadings for prior wakefulness 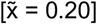; factor 2 had high loadings for sleep staging (first deep sleep segment length 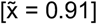 and amount of deep sleep before first REM segment 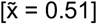). The final S_b_ score emphasized the first deep sleep segment [0.32], followed by steps [0.24], activity intensity [0.2], and deep sleep prior to first REM segment [0.28] (**Fig 2G**). Prior wakefulness was de-emphasized in the final score [0.06].

### Interpreting C_b_ and S_b_ scores

These scores are oriented so that lower scores reflect better behaviors for both processes. They can be interpreted at the day-level (e.g., Tuesday had a high C_b_ score because the participant went to bed very late, or Wednesday had a low S_b_ score because the participant was active and did not nap) or at the aggregate level (e.g., participant X has a lower mean C_b_ score).

### Assessing consistency of behavior scores in situations known to affect the two-process model

To characterize our C_b_ and S_b_ scores in the context of behaviors related to the expected underlying neurobiological processes, we examined them in contexts where we would expect to see certain patterns.

#### Process C

Process C is known to vary with age - middle-age to older individuals tend to sleep and wake earlier and have more stable sleep schedules compared to adolescents and young adults.^17–19^ Mirroring this relationship, our C_b_ scores showed that C_b_ generally decrease as age increases (**Figure 4A**). Notably, there appeared to be a stabilization across scores in middle age, and then a further decrease in much older age.

**Figure 4.**
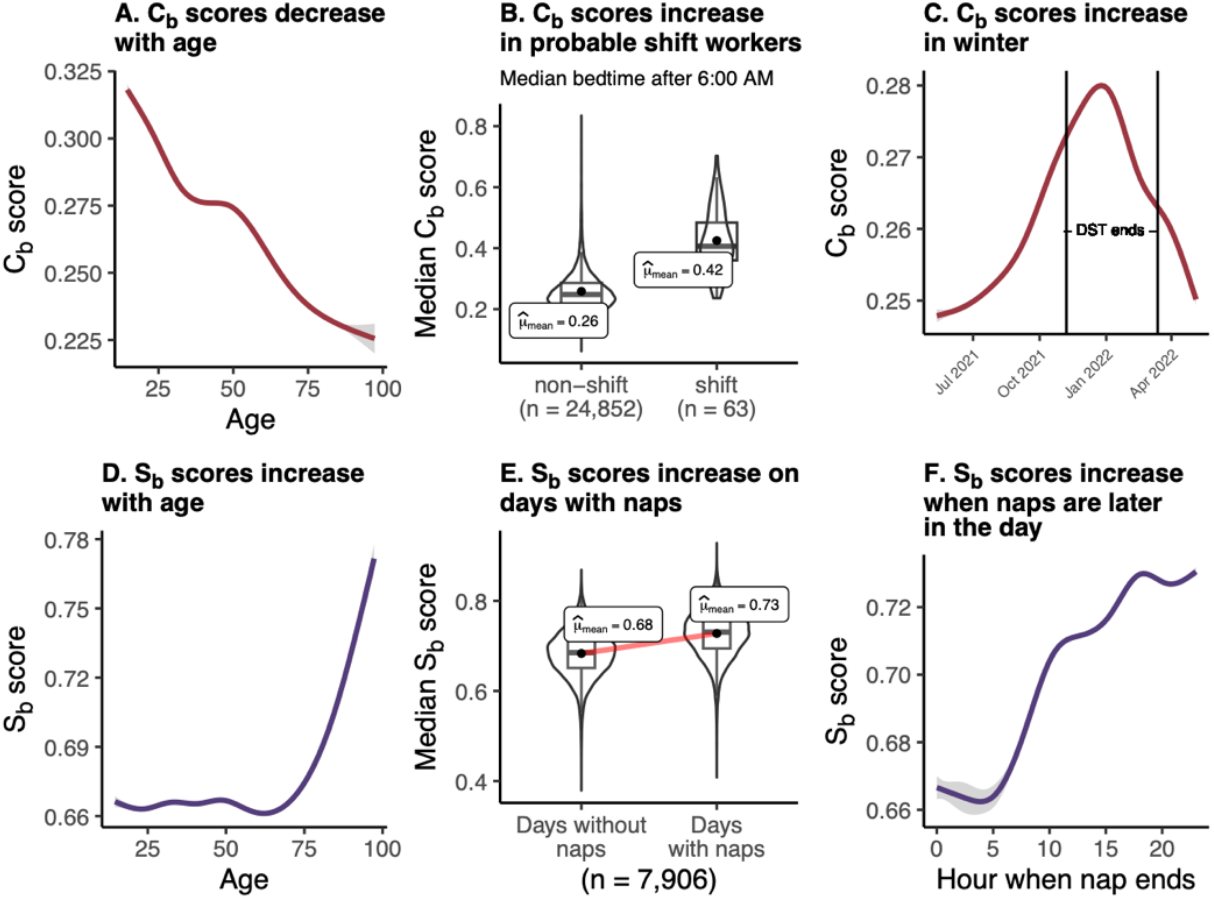
Consistency of C_b_ and S_b_ scores in applied scenarios. Lower scores reflect better behaviors. Line plots show smoothed mean composite scores with confidence intervals across our 27,313 individuals to examine overall trends in our data. Age is the age at each day. **(A)** C_b_ scores decrease linearly as age increases, while **(D)** S_b_ scores increase exponentially at the end of life. **(B)** C_b_ scores are higher in participants who are likely shift workers (n = 63), defined as having a median bedtime later than 6 AM. **(C)** C_b_ scores increase during Winter, exemplified here with data from 2021-2022. **(E)** In 7,906 individuals with at least 10 naps, median S_b_ scores were increased on days with naps. **(F)** S_b_ scores are further increased the later those naps are in the day.

**Figure 5.**
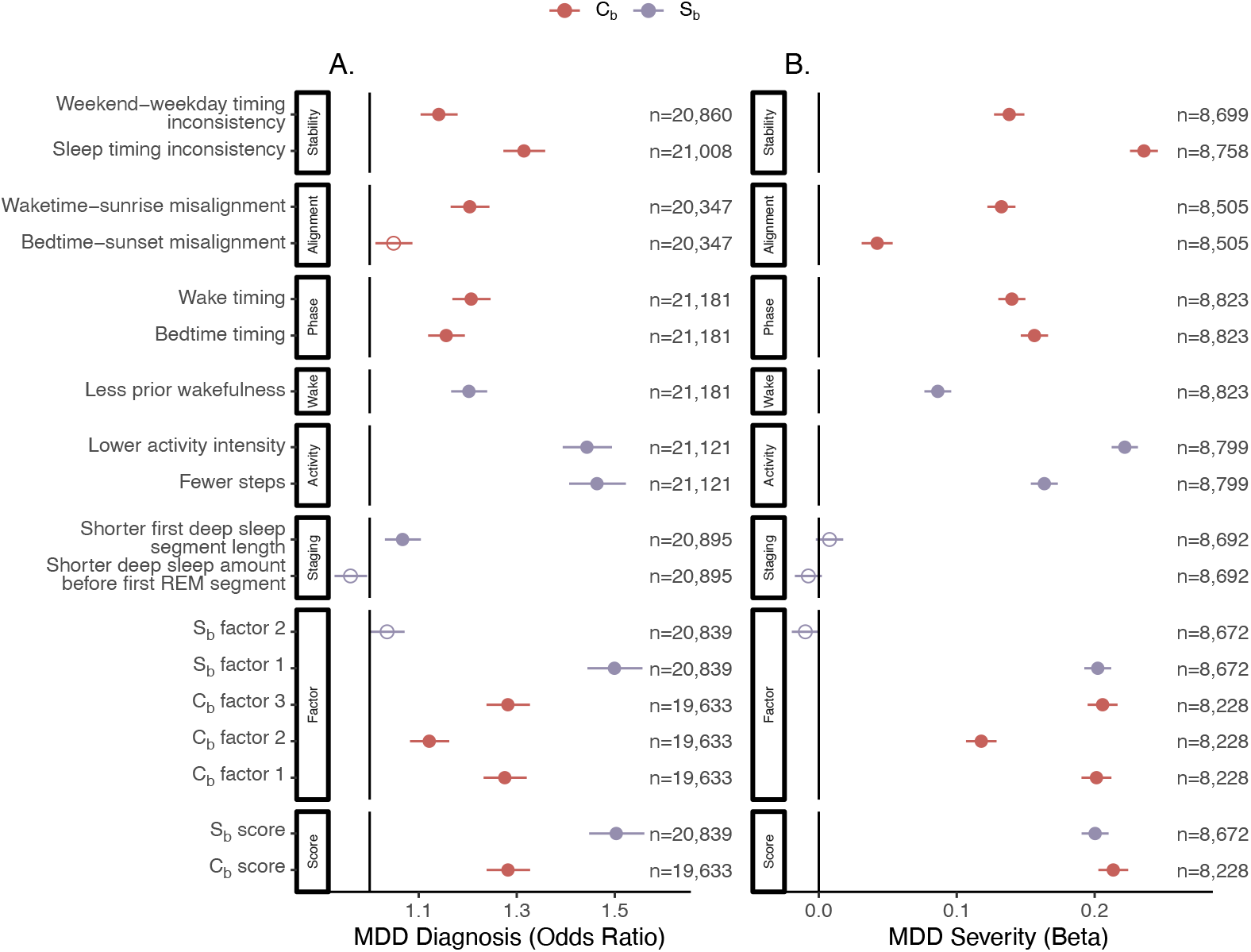
Association of C_b_ and S_b_ and scores with depression. Results of regression analysis of C_b_ and S_b_ and scores, covarying for age, gender, race, and Fitbit observation length. **(A)** Association (odds ratio) of behaviors and scores with Major Depressive Disorder (MDD) diagnosis, defined by electronic heath records. **(B)** Association (beta) of behaviors and scores with depression severity, measured by Patient Health Questionnaire-9 (PHQ-9).

Next, we sought to see whether C_b_ scores were increased in those with constrained sleep schedules, like shift workers.^20^ We identified 63 individuals in our cohort who have a median bedtime after 6:00 AM, and found these individuals had significantly higher C_b_ scores, indicating worse C_b_ (**Figure 4B**).

Finally, we expect to see higher C_b_ scores in the winter compared to the summer. This relationship is expected because earlier sunsets in the winter decrease sunset-bedtime alignment, and lower sunlight exposure has large effects on the circadian melatonin system.^21^ We therefore examined C_b_ scores across seasons (**Figure 4C**). As expected, C_b_ scores were higher during the winter and lower during the summer.

#### Process S

Process S is known to deteriorate with age, explained by a reduction of deep sleep and increased sleep fragmentation.^22^ Reflecting this, our S_b_ scores were strongly associated with age, notably in much older adults (>70) (**Figure 4D**).

An integral finding of the original two-process model was that naps greatly decrease process S buildup, as they relieve the pressure built up during wakefulness if the nap contains deep sleep.^8,23^ We do not include naps directly as a behavior in our scores, and instead use the amount of prior wakefulness leading up to the sleep period (either wakefulness from waking up to bedtime or from the end of a nap to bedtime). Here, we can see that S_b_ scores were significantly higher on days with naps, as expected (**Figure 4E**). S_b_ scores were also higher on days with naps later in the day (**Figure 4F**), reflecting the need for wakefulness prior to sleep to build sleep pressure.

### Representation of C_b_ and S_b_ scores in depression

To help interpret the scores, we examined them in individuals with depression, since depression was the primary disorder used to untangle the two-process model and is expected to have relationships with both process C and S.^8^ We modeled depression diagnosis risk using logistic regression (controlling for age, gender, race, and number of Fitbit observations) and median value of each behavior and score. We required an individual to have at least 14 observations of the predictor behavior or score to be included in each regression. We found that S_b_ scores were much higher in participants with an MDD diagnosis, indicating that higher S_b_ scores are associated with more risk of MDD (OR = 1.5, CI [1.44, 1.56], p < 2e-16). This relationship appears to be mostly driven by activity features (steps OR =1.46, CI [1.41, 1.53], p < 2e-16; steps intensity OR = 1.44, CI [1.4, 1.5], p < 2e-16), though prior wakefulness (OR = 1.2, CI [1.17, 1.24], p < 2e-16) and shorter initial deep sleep segments (OR = 1.07, CI [1.03, 1.1], p = 0.0002) were also significantly associated with diagnosis. C_b_ scores were also associated with diagnosis, though at a lower effect size (OR = 1.28, CI [1.24, 1.33], p < 2e-16).

We then examined whether C_b_ and S_b_ scores were related to depression severity in a subset of 10,758 individuals who completed a PHQ-9 survey. Survey score was used as a severity metric and tested in the same linear regression as described above. Notably, C_b_ and S_b_ scores were similarly associated with depression severity (S_b_ score β = 0.2, 95% CI [0.196, 0.204], p < 2e-16; C_b_ score β = 0.21, 95% CI [0.209, 0.218], p < 2e-16). C_b_, specifically bedtime timing (β = 0.156, 95% CI [0.153, 0.159], p < 2e-16) and sleep timing inconsistency (β = 0.236, 95% CI [0.23, 0.24], p < 2e-16), displayed much higher effect sizes with depression severity than diagnosis.

## Discussion

To improve interpretation of wearable derived sleep data, we constructed scores based on behaviors tied to the neurobiological understanding of sleep in the two-process model. By separating Fitbit behaviors into process C (circadian) and S (homeostatic) and examining how those features interact at the level of a single day, we can identify which sleep regulation processes are impaired, by how much they are impaired, and what individual behavioral modifications could be performed to improve health outcomes. For example, a person with a low process S_b_ score over several to many days may not be getting enough deep sleep which could be modified with changes in bed or wake time or by increasing activity. This detailed understanding of behaviors tied to neurobiology provides an opportunity to recommend individual-level changes to improve sleep and overall health.

C_b_ and S_b_ scores should be interpreted as behaviors related to underlying process C and S, allowing the concepts of circadian and homeostatic sleep processes to be studied at scale for the first time. While C and S themselves can be calculated mathematically from polysomnography and EEG,^24^ these measurement systems are not suited for large sample sizes or longitudinal monitoring. We expect our scores to be correlated with individual differences in actual C and S and have shown that they behave as expected in real-world scenarios, such as finding C_b_ disruptions in sleep workers and S_b_ disruptions on days with naps. The advantage of these behavioral scores is the identification of longitudinal patterns among large sample sizes, which then present opportunities for behavioral modification, impacting the underlying trait-like C and S.

When interpreting our scores by relating them to depression, we found that while process S_b_ are most correlated with depression diagnosis, C_b_ and S_b_ are equally associated with depression severity. This finding implies that S_b_ may be more indicative of likelihood to develop depression, but that both impact the severity of that depression. There is a known strong association of process S with depression known as the S-deficiency hypothesis, which posits that sleep issues in depression are caused by a lack of S buildup throughout the day as a result of low activity and disruption of serotonergic systems.^8,9^ Interestingly, we see that C_b_ and S_b_ appear to be equally impactful on depression severity regardless of diagnosis. This could be because dysregulation of S eventually leads to dysregulation of C, or because individuals with poor C_b_ that also have depression have worse outcomes. This finding invites further study of the impact of C_b_ and S_b_ on depression symptoms and severity to find optimal behavioral interventions.

A limitation of this work is the unreliability of Fitbit sleep staging data.^25^ Sleep staging is the classical way of numerically defining process S, specifically through REM latency (the amount of time between sleep onset and the first REM cycle).^5,6^ However, we found REM latency to be very unreliable in our cohort **(Supplemental Fig 4)**, likely due to Fitbit’s overestimation of REM sleep and underestimation of deep sleep.^26^ To overcome this, we examined many relevant staging features to see if any followed the neurobiologically expected patterns of covariance, specifically where less wakefulness and more napping should have a negative impact on S-related sleep features. We identified two deep sleep related features that followed the expected direction, but even these were only weakly correlated in the right directions **(Supplemental Fig 4)**. In future work, if staging data from wearables becomes more reliable, this should be re-evaluated, as REM latency is likely to be more informative. Additionally, while lower scores reflect generally “better” behaviors, the assumptions made should still be examined in context. For example, we know that process C deterioration occurs in the elderly in the form of reduced circadian arousal in the evening, which may be reflected in our scores as earlier sleep timing.^17,22,27^

In summary, this work attempts to make Fitbit sleep behaviors more interpretable for intervention and use in future research. We have utilized biological frameworks to inform Fitbit interpretation and thoroughly examined the credibility and usefulness of this framework. This work creates many further opportunities to examine sleep in health contexts, including longitudinal studies of behavioral changes surrounding treatment or diagnosis, association of sleep behaviors with disease, and individualized recommendations for changes in sleep behaviors to improve health.

## Data Availability

All data produced in the present study are available upon reasonable request to the authors

## Acknowledgements

The All of Us Research Program would not be possible without the partnership of its participants. The All of Us Research Program is supported by the National Institutes of Health, Office of the Director: Regional Medical Centers (1 OT2 OD026549; 1 OT2 OD026554; 1 OT2 OD026557; 1 OT2 OD026556; 1 OT2 OD026550; 1 OT2 OD 026552; 1 OT2 OD026553; 1 OT2 OD026548; 1 OT2 OD026551; 1 OT2 OD026555; IAA: AOD21037, AOD22003, AOD16037, AOD21041), Federally Qualified Health Centers (HHSN 263201600085U), Data and Research Center (5 U2C OD023196), Biobank (1 U24 OD023121), The Participant Center (U24 OD023176), Participant Technology Systems Center (1 U24 OD023163), Communications and Engagement (3 OT2 OD023205; 3 OT2 OD023206) and Community Partners (1 OT2 OD025277; 3 OT2 OD025315; 1 OT2 OD025337; 1 OT2 OD025276).

