## Supplemental Figures for "Making sleep behaviors interpretable: adapting the two-process model of sleep regulation to longitudinal Fitbit sleep and activity behaviors for health insights"

S1. Distributions of C_b_ and S_b_ across BYOD and WEAR sub-study cohorts.

S2. Quality control pipeline for Fitbit data.

S3. Schematic of C_b_ and S_b_ at the day-level.

S4. Selection of sleep staging features for process S.

S5. Correlation of prior wakefulness with other process S features by amount of confirmed wear time.

S6. Group membership of Major Depressive Disorder (MDD) diagnosis criteria.

S7. Scree plots of S_b_ and C_b_ to determine number of factors for exploratory factor analysis

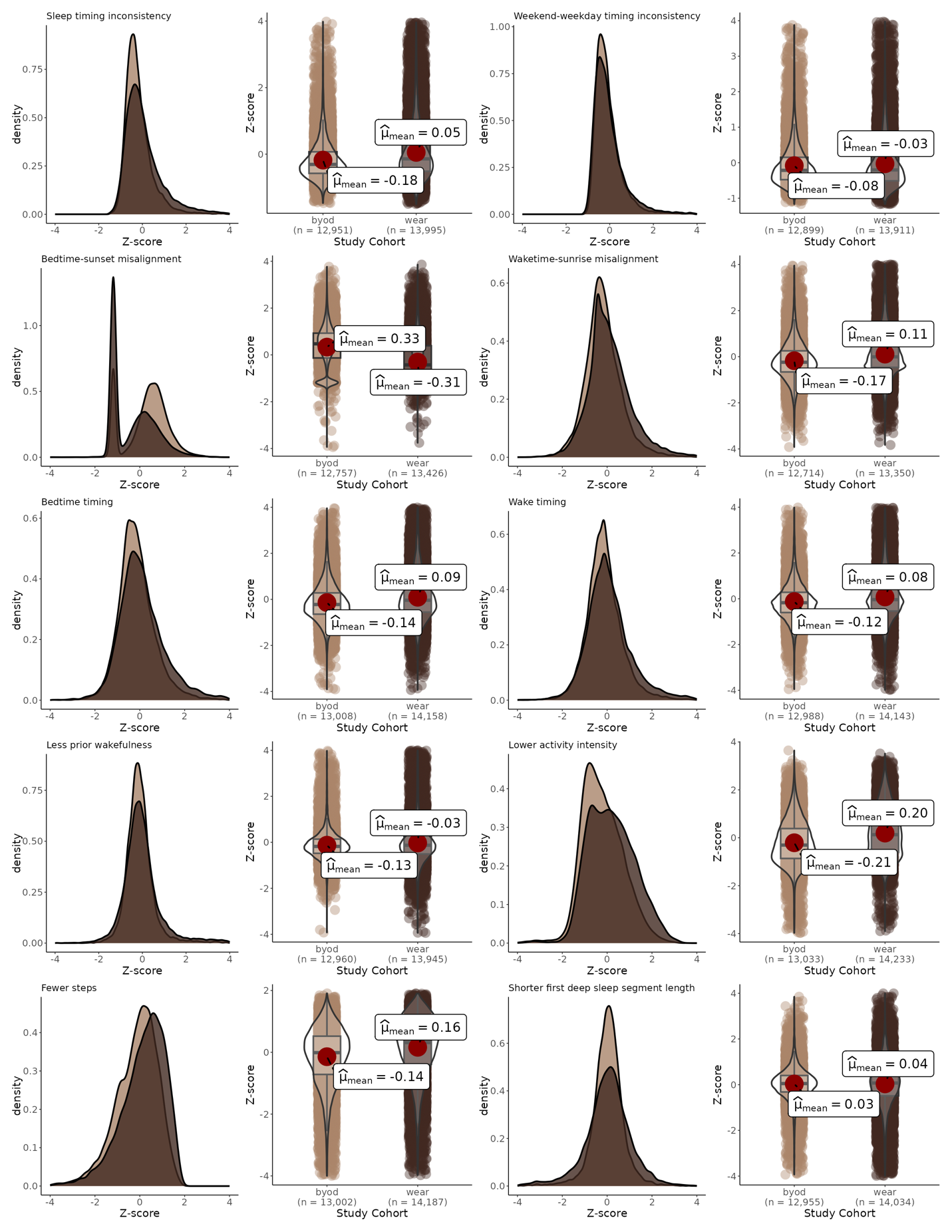

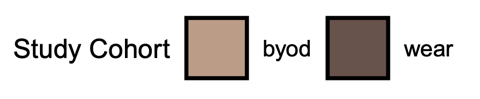

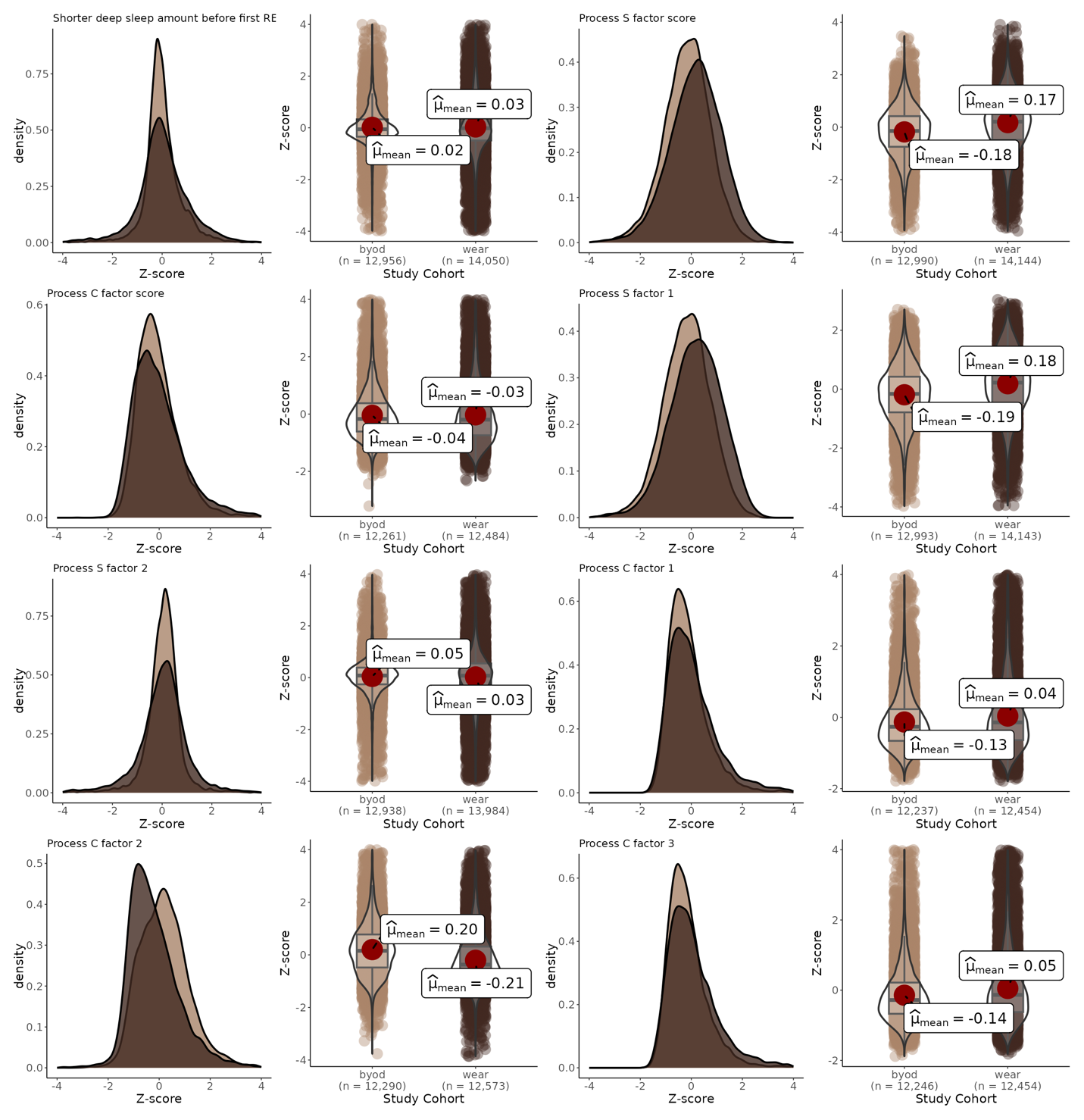

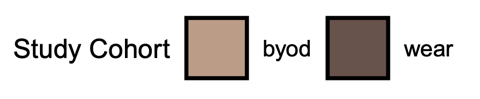

*S1. Distributions of behaviors and scores across bring-your-own-device (BYOD) and WEAR study sub-cohorts.*

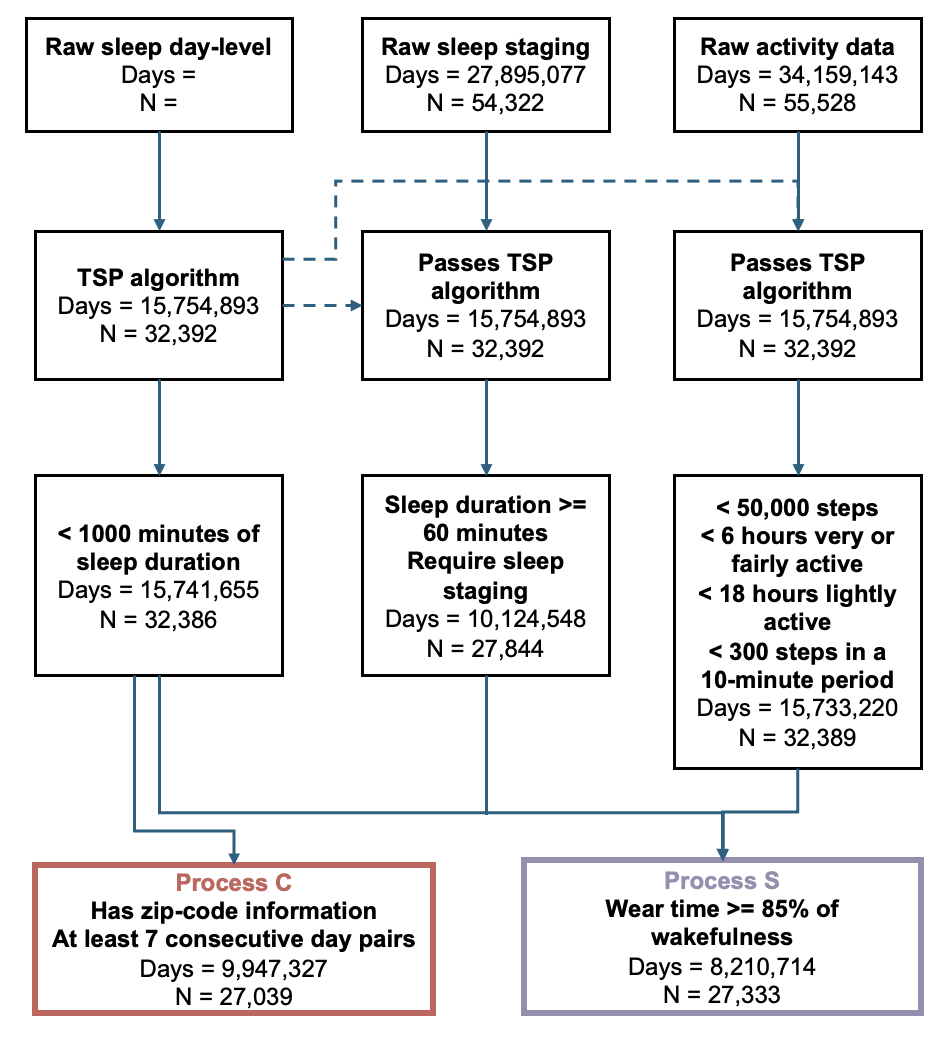

*S2. Quality control pipeline for Fitbit data.*

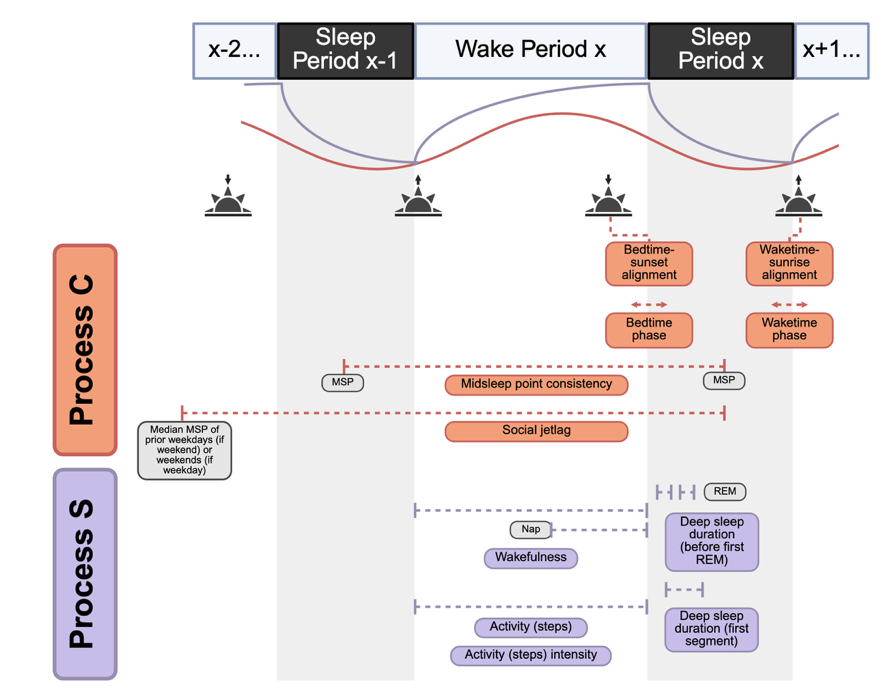
*S3*. *Schematic of C_b_ and S_b_ at the day-level.*

Features were calculated at the level of each day, so that every day had a set of C_b_ and S_b_. For each main sleep period X, data were incorporated from that night, the prior waking hours, the prior night, and the nights in the weekend or weekday prior.

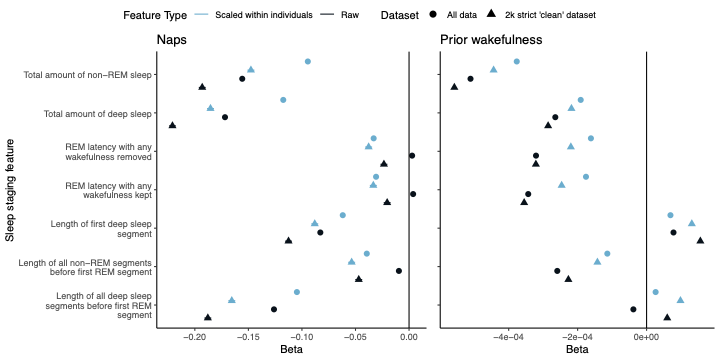

*S4. Selection of sleep staging features for process S.*

In our dataset, we found that Fitbit sleep staging features were not associated with process S behaviors at the day-level in the expected directions based on the neurobiology of sleep regulation. Namely, days without naps and with longer prior wakefulness should be associated with increased REM latency and increased deep sleep. It was unclear whether this lack of the expected association was due to Fitbit measurement differences or whether it was an actual finding. To explore this, we engineered seven sleep staging phenotypes that should be associated negatively with naps and positively with wakefulness. We examined these staging features in the entire dataset and in a strict “healthy” subset of 2,453 individuals with no history of MDD, insomnia, sleep apnea, or an antidepressant, who were younger than 50 and had at least 100 valid Fitbit days, and who had a median bedtime between 8pm-2am and a median waketime between 5-11am. Additionally, due to high individual differences in sleep staging, we also examined these sleep features scaled within each individual. We then ran linear regressions controlling for age at entry, gender, race, and number of Fitbit observations.

We identified two staging features that exhibited the correct directional association with naps and wakefulness: the length of the first deep sleep segment, and the length of all deep sleep segments prior to the first REM segment. We also found that scaling within the individual increased the association between these features and naps and wakefulness and chose to use them in subsequent analysis as process S-related sleep staging features.

As for why some of the expected Fitbit features do not have the expected relationships, it is possible that the underestimation of deep sleep and overestimation of REM sleep in Fitbit devices is confounding the relationships we expect to see. We believe that the two features selected are representative of the underlying process S neurobiology in Fitbit devices.

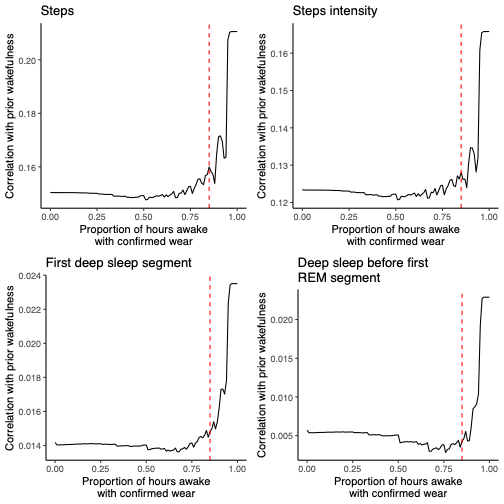

*S5. Correlation of prior wakefulness with other process S features by amount of confirmed wear time.* As the amount of confirmed wear time increased, the correlation of prior wakefulness with other process S features increased, indicating that wakefulness without confirmed wear is not reliable in our cohort. We therefore added a criteria to require at least 85% of wakefulness before sleep to have confirmed wear time, indicated here by the dashed red line.

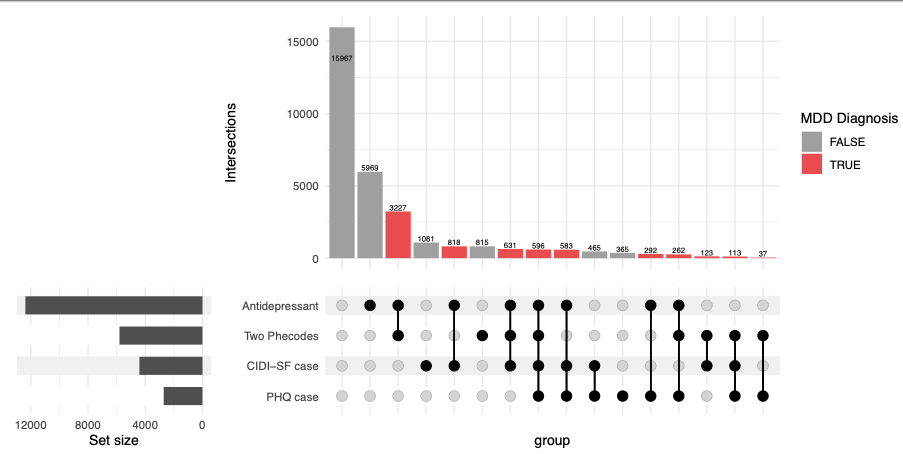

*S6. Group membership of Major Depressive Disorder (MDD) diagnosis criteria.*

To be considered a diagnosed case of MDD, an individual had to meet at least two of the three
following criteria: having at least two instances of an MDD phecode on different dates; one prescription for an antidepressant; a score of >10 on the PHQ-9 or qualifying for case status on the CIDI-SF, when available.

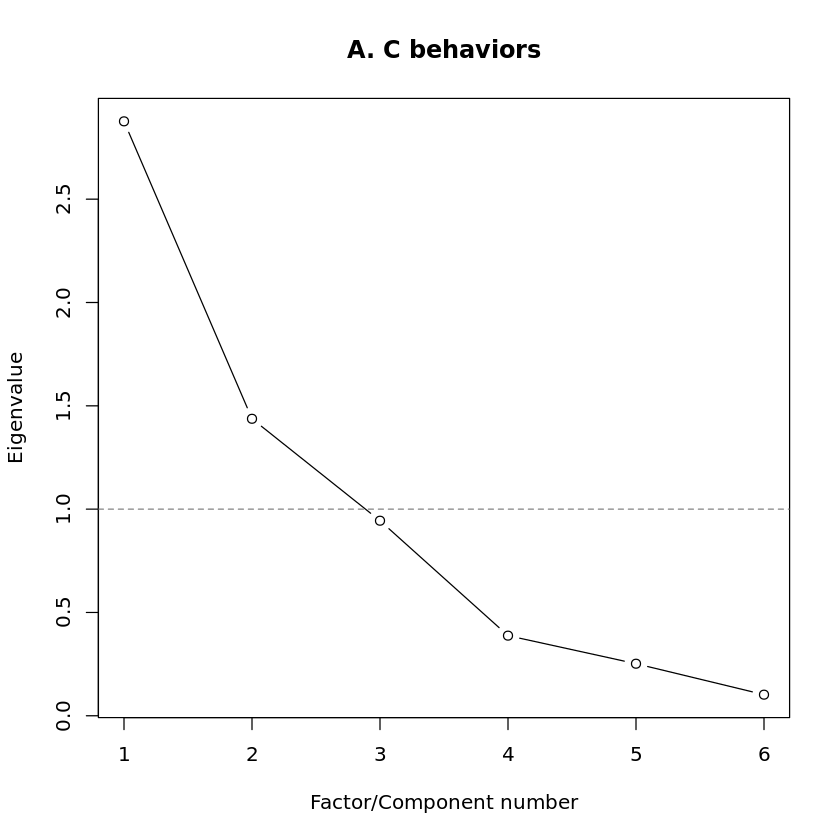

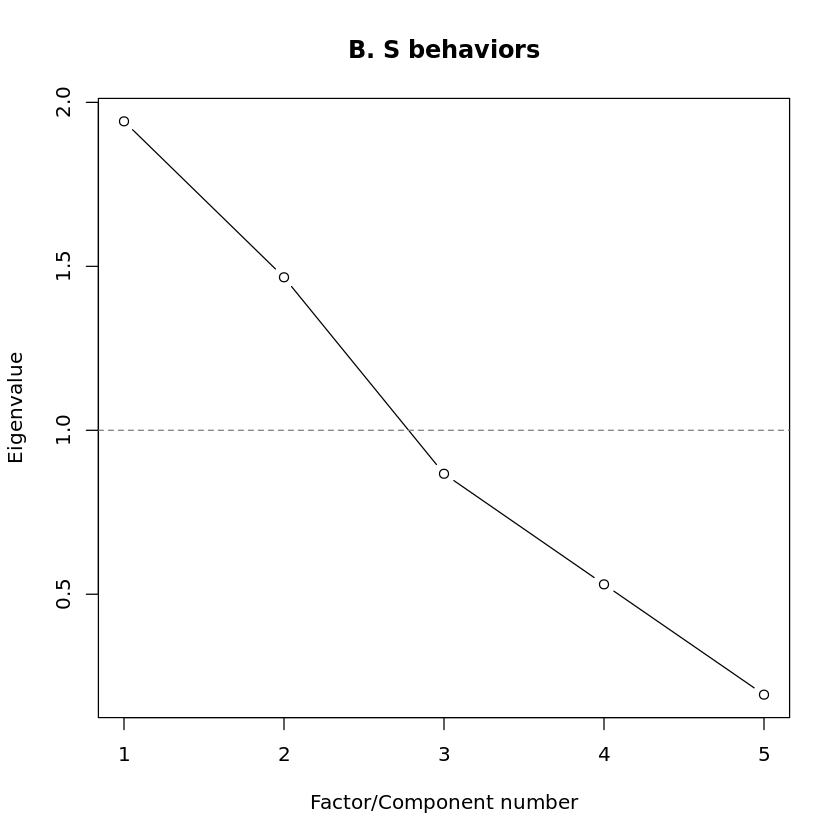

*S7. Scree plots of S_b_ and C_b_ to determine number of factors for exploratory factor analysis.*
